# The association of infectious mononucleosis and breast cancer in The Health of Women (HOW) Study®

**DOI:** 10.1101/2021.10.05.21264564

**Authors:** Yujing J. Heng, Susan Love, Jessica Clague DeHart, Joyce D. Fingeroth, Gerburg M. Wulf

## Abstract

**Background:** The link between Epstein-Barr Virus (EBV) and breast cancer (BC) remains unclear. Infectious mononucleosis (IM) is a clinical manifestation of delayed onset of EBV infection in early adulthood. We utilized the Health of Women (HOW) Study® to understand the association between IM and BC risk.

**Subjects and methods:** The HOW Study® was a web-based survey of BC risk factors with >40,000 participants who answered seven modules between 2012 and 2015; 3,654 women had IM between the ages of 10 and 22 years (16.8%) and 17,026 never developed IM (78.5%). Of these 20,680 women, 1,997 (9.7%) had Stages I-III BC and 13,515 (65.4%) were cancer-free. Multivariable binary logistic regression ascertained the association between IM and BC risk by controlling for ethnicity, family history, age at menarche, oral contraceptive use, tobacco use, birthplace, parity, age at first birth, body mass index, and breast biopsy. Secondary analyses stratified cancer cases into those who had BC at <50 or ≥50 years old and by estrogen receptor (ER) subtype.

**Results:** Participants were mostly white, middle-aged women born in the United States or Canada. Women who had IM were less likely to develop BC than those who did not develop IM (adjusted odds ratio (OR)=0.83, 95% confidence interval (CI) 0.71-0.96). Findings were similar when stratifying women into <50 or ≥50 years old at BC diagnosis (<50 years old, adjusted OR=0.82, 95% CI 0.67-0.998; ≥50 years old, adjusted OR=0.83, 95% CI 0.69-1.00). Women who had IM were less likely to develop ER positive BC (adjusted OR=0.84, 95% CI 0.71-0.997); there was no association between IM and ER negative BC (adjusted OR=0.88, 95% CI 0.65-1.16).

**Conclusion:** In the HOW Study®, women diagnosed with IM between the ages of 10 and 22 had lower breast cancer risk compared to women who never developed IM.

## Introduction

Over ninety percent of the world’s adult population are exposed to Epstein-Barr Virus (EBV) during their childhood [1]. Early life EBV infections typically manifest as subclinical illness whilst delayed onset into early adulthood may manifest as infectious mononucleosis (IM) [2]. EBV infection is an established risk factor in the subsequent development of Hodgkin’s lymphoma [3], African Burkitt’s lymphoma [4], gastric cancer [5], and nasopharyngeal carcinoma [6]. The ubiquitous nature of EBV infection and its link to cancer has prompted a call for an EBV vaccine [7,8].

In 1995, Labrecque *et al*. published the first report about EBV in breast tumors [9]. Since then, the majority of research in this area was geared towards detecting and describing the frequency of EBV positive breast tumors [10–19]. Our group [20] and others [21,22] have conducted preclinical *in vivo* and *in vitro* studies to understand the mechanisms of EBV in breast cancer cells. Unsurprisingly, the reports regarding the frequencies of EBV positive breast cancer in pathology specimens are inconsistent and may be explained by differences in geography and laboratory methods used to detect this herpesvirus—polymerase chain reaction [10–12,18,19,23–28], immunostaining for EBV-encoded nuclear antigen 1/2 [10,12–14,25,26], or *in situ* hybridization for small EBV-encoded RNAs [12,15–17]. EBV positive breast tumors are less frequent in the United States (0-25%) [12,15,16], Mexico (0-5%) [28], Germany (7%) [13], and Iran (0%) [26], compared to Argentina (35%) [23], United Kingdom (21%) [9,10], France (27-51%) [18,29], The Netherlands (33%) [29], Denmark (35%) [29], Portugal (25%) [24], Algeria (40%) [29], Tunisia (27-33%) [29,30], Eritrea (28-36%) [19], Egypt (45%) [31], Iraq (28%) [31], Lebanon (40%) [25], India (30-55%) [14,17], Pakistan (24%) [27], and China (60%) [11]. However, three meta-analyses conducted using these histopathology-based studies listed above concluded that EBV increases breast cancer risk [32–34].

The life-long association of EBV in a large population of people makes it challenging to clarify a causal link between this virus and breast cancer [35]. When IM is used as a surrogate for latent EBV infection, epidemiological investigations do not support an association between EBV infection and breast cancer risk [36–38]. For example, we previously reported no association between IM and breast cancer risk using data from the Nurses’ Health Study II, a prospective study of >100,000 young female nurses in the United States [37]. Further investigations into the association between IM and breast cancer risk is warranted in order to understand whether IM may be a breast cancer risk factor, as well as inform the usefulness of an EBV vaccine as a breast cancer prevention tool. In this current study, we utilized The Health of Women (HOW) Study® to further explore the association between IM and breast cancer risk.

## Materials and methods

### Study Population

The HOW Study® (NCT02334085) was a collection of cross-sectional, web-based surveys of breast cancer risk factors completed by participants aged 18 or older with and without breast cancer [39]. Most participants were residing in the United States at the time of survey (<1% international responses) [39]. The HOW Study® consisted of seven modules released sequentially from 2012-2015: 1) *My Health Overview*, 2) *My Breast Cancer*, 3) *My Personal and Family Health History*, 4) *Health, Weight, and Exercise*, 5) *Environmental Exposure*, 6) *Quality of Life*, and 7) *Bacteria in the Breast*. These modules assessed health histories (general, reproductive, and family), lifestyle factors, environmental exposures, breast cancer diagnosis and treatment, and cancer survivorship life quality. Survey enrollment closed in 2019; data in this manuscript were from the 2016 data freeze. Participants provided written consent. The HOW Study® protocol was approved by the Western Institutional Review Board.

### Ascertainment of IM

*My Health Overview* collected information related to socio-demographics, current health status and behaviors, reproductive history, and data on health-limiting activities. It was completed by 42,540 participants with 13,285 (31.2%) indicating they had breast cancer [39]. A subset of female participants between the ages 18 and 91 also completed *My Personal and Family Health History* (*n*=22,355 out of 42,540; 52.6%). These women identified as cisgender female on both modules. *My Personal and Family Health History* collected information about genetic risk factors or predispositions for breast cancer, including whether they had IM (yes/no; *n*=21,701). Women were excluded if they indicated “don’t know” or if they did not answer the question (*n*=654; 2.9%).

If the woman answered “yes”, they were asked for their age at the time of IM diagnosis (<10 years old, 10-22 years old, >22 years old, or unknown) and how IM was diagnosed (by physician based on symptoms, laboratory blood test, lymph node biopsy, or unknown). We focused on women who had IM between 10 and 22 years old (*n*=3,654; 16.8%) and those who indicated they never developed IM (*n*=17,026; 78.5%). Women who had IM at <10 years old (*n*=183; 0.8%), >22 years old (*n*=764; 3.5%), or unknown age (*n*=74; 0.3%) were excluded from this study due to insufficient power.

### Ascertainment of primary breast cancer

The ascertainment of first primary breast cancer and age at diagnosis were derived from *My Health Overview, My Breast Cancer, Quality of Life*, and *Bacteria in the Breast*. Of the remaining 20,680 women who either had IM between the ages of 10 and 22 or never developed IM, 1,997 (9.7%) had Stages I-III breast cancer and 13,515 (65.4%) were breast cancer-free. We excluded 1,398 (6.8%) who self-reported *in situ* breast cancer, 45 (0.2%) with Stage IV, 3,707 (17.9%) who had breast cancer but it was unclear whether the cancer was *in situ* or invasive, and 18 (0.1%) with unknown breast cancer status. Age at invasive breast cancer diagnosis was grouped into 20-24, 25-29, 30-34, 35-39, 40-44, 45-49, 50-54, 55-59, 60-64, 65-69, 70-74, 75-79, and ≥80 years old. No breast cancer case occurred before the onset of IM.

### Statistical analysis

We determined whether IM was associated with later development of invasive breast cancer by using multivariable binary logistic regression to calculate odds ratios (ORs) and 95% confidence intervals (CIs). The HOW Study® modules did not specifically collect epidemiological data closest to or at time of breast cancer diagnosis. In model 1, we controlled for potential confounding of these variables that were likely to be similar at time of breast cancer diagnosis and at the time of the survey: ethnicity (white versus others), family history of female breast cancer (yes/no), age at menarche (<10, 10, 11, 12, 13, 14, 15, 16, or ≥17 years old), use of oral contraceptives (ever/never), any tobacco (ever/never), and birthplace (United States/Canada versus foreign born). We decided to include birthplace because subclinical EBV infection may be acquired in childhood.

For model 2, we controlled for variables in model 1 as well as these additional variables obtained at time of survey that may not accurately reflect the status at time of breast cancer diagnosis: parity (nulliparous, primiparous, multiparous, or unknown), age at first birth (<18, 18-24, 25-29, 30-34, 35-39, 40-44, or ≥45), BMI (kg/m^2^), and had a breast biopsy (ever/never). These variables were extracted from *My Health Overview* and *My Personal and Family Health History*. We conducted sensitivity analyses by restricting to women born in the United States or Canada as well as women diagnosed with IM using a laboratory blood test. Finally, we conducted secondary analyses by stratifying cancer cases into those who had breast cancer at <50 or ≥50 years old; and by estrogen receptor (ER) status. We were unable to evaluate menopausal status at time of first primary breast cancer (>66.7% missing data) or at time of survey (>40% missing data).

## Results

Our study participants consisted of 2,773 (17.9%) women who had IM between the ages of 10 and 22 and 12,739 (82.1%) who did not have IM. The majority of women who had IM were diagnosed with a laboratory blood test (53.1%; Table 1). Our women were mostly white, middle-aged, and born in the United States or Canada (Table 1). Among the 80 women who had IM and were not born in the United States or Canada, 63 (78.8%) indicated they have been living in the United States for >10 years, 1 (1.2%) woman has been living in the United States between 4 to 7 years, and 16 (20.0%) did not answer. The variables of interest were similar between those who had IM and did not develop IM (Table 1). Most women had a family history of female breast cancer (>50%), never had a breast biopsy, started menstruating around 12 years old, were multiparous, had normal BMI, used oral contraceptives, but were less likely to use tobacco. Women who had IM were more likely to have breast fed for at least 12 months.

**Table 1.**
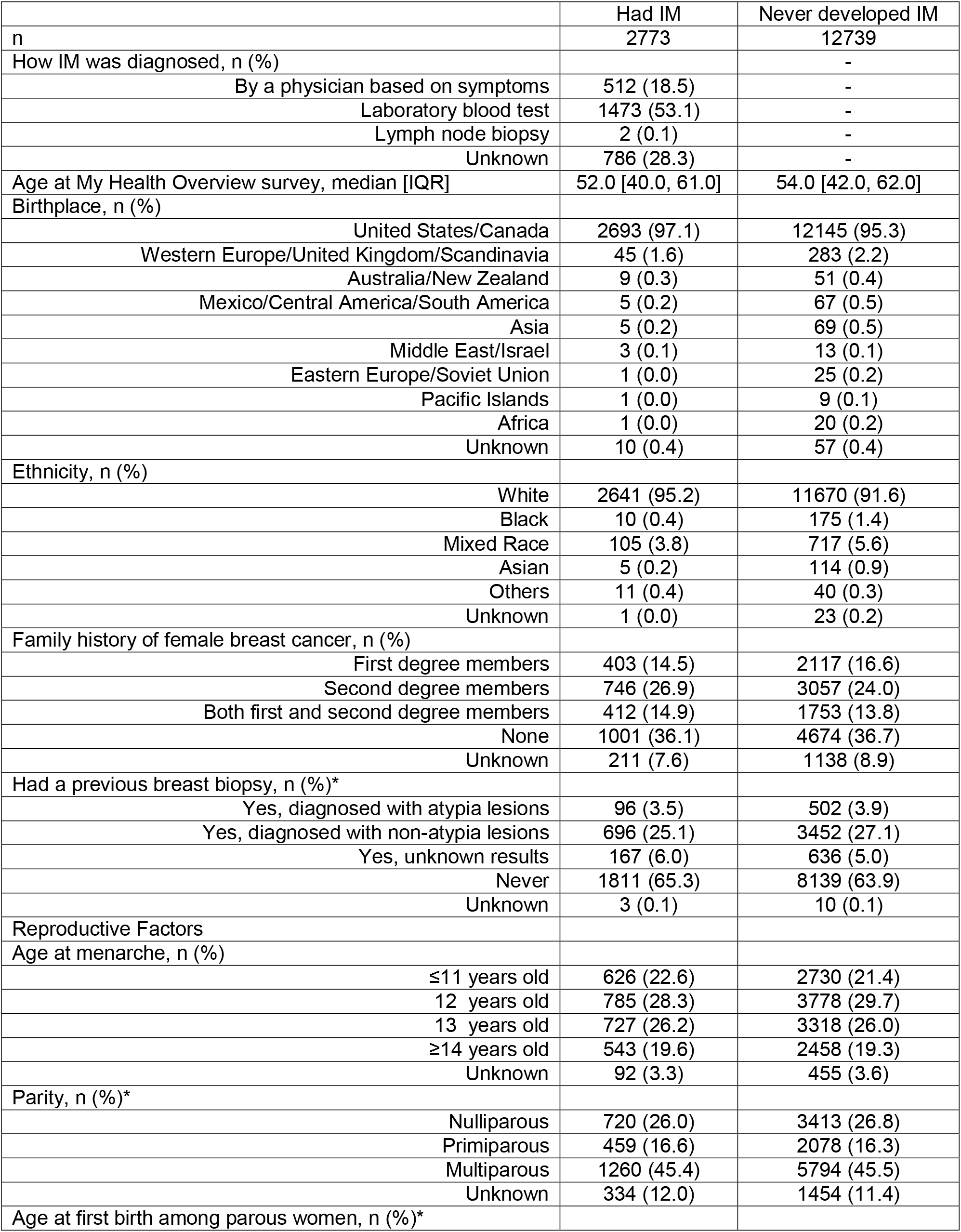
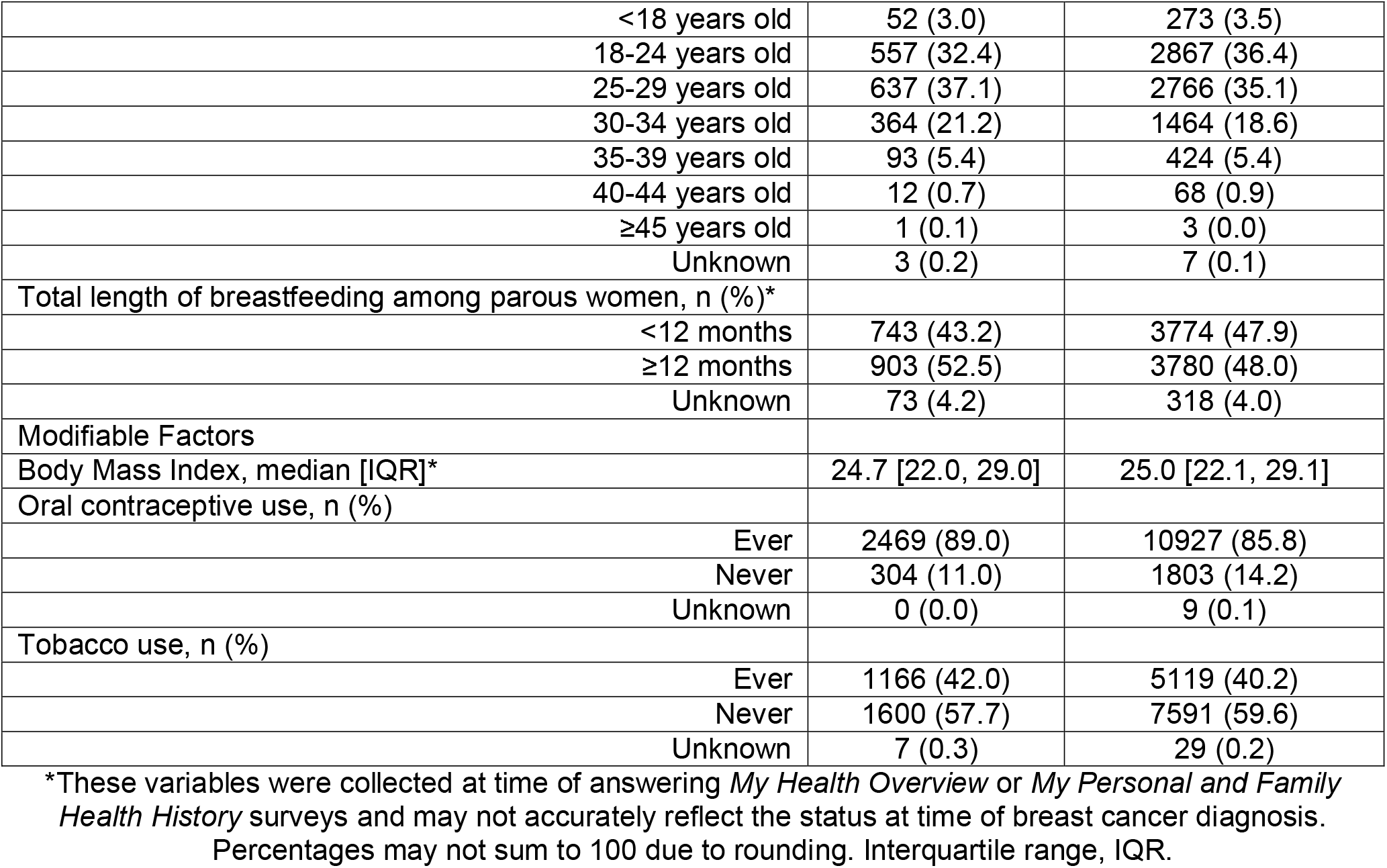
Demographic and breast cancer risk factors of study participants who had infectious mononucleosis (IM) between ages 10 and 22 and those who never developed IM.

|  | Had IM | Never developed IM |
| --- | --- | --- |
| n | 2773 | 12739 |
| How IM was diagnosed, n (%) |  | - |
| By a physician based on symptoms | 512 (18.5) | - |
| Laboratory blood test | 1473 (53.1) | - |
| Lymph node biopsy | 2 (0.1) | - |
| Unknown | 786 (28.3) | - |
| Age at My Health Overview survey, median [IQR] | 52.0 [40.0, 61.0] | 54.0 [42.0, 62.0] |
| Birthplace, n (%) |  |  |
| United States/Canada | 2693 (97.1) | 12145 (95.3) |
| Western Europe/United Kingdom/Scandinavia | 45 (1.6) | 283 (2.2) |
| Australia/New Zealand | 9 (0.3) | 51 (0.4) |
| Mexico/Central America/South America | 5 (0.2) | 67 (0.5) |
| Asia | 5 (0.2) | 69 (0.5) |
| Middle East/Israel | 3 (0.1) | 13 (0.1) |
| Eastern Europe/Soviet Union | 1 (0.0) | 25 (0.2) |
| Pacific Islands | 1 (0.0) | 9 (0.1) |
| Africa | 1 (0.0) | 20 (0.2) |
| Unknown | 10 (0.4) | 57 (0.4) |
| Ethnicity, n (%) |  |  |
| White | 2641 (95.2) | 11670 (91.6) |
| Black | 10 (0.4) | 175 (1.4) |
| Mixed Race | 105 (3.8) | 717 (5.6) |
| Asian | 5 (0.2) | 114 (0.9) |
| Others | 11 (0.4) | 40 (0.3) |
| Unknown | 1 (0.0) | 23 (0.2) |
| Family history of female breast cancer, n (%) |  |  |
| First degree members | 403 (14.5) | 2117 (16.6) |
| Second degree members | 746 (26.9) | 3057 (24.0) |
| Both first and second degree members | 412 (14.9) | 1753 (13.8) |
| None | 1001 (36.1) | 4674 (36.7) |
| Unknown | 211 (7.6) | 1138 (8.9) |
| Had a previous breast biopsy, n (%)* |  |  |
| Yes, diagnosed with atypia lesions | 96 (3.5) | 502 (3.9) |
| Yes, diagnosed with non-atypia lesions | 696 (25.1) | 3452 (27.1) |
| Yes, unknown results | 167 (6.0) | 636 (5.0) |
| Never | 1811 (65.3) | 8139 (63.9) |
| Unknown | 3 (0.1) | 10 (0.1) |
| Reproductive Factors |  |  |
| Age at menarche, n (%) |  |  |
| ≤11 years old | 626 (22.6) | 2730 (21.4) |
| 12 years old | 785 (28.3) | 3778 (29.7) |
| 13 years old | 727 (26.2) | 3318 (26.0) |
| ≥14 years old | 543 (19.6) | 2458 (19.3) |
| Unknown | 92 (3.3) | 455 (3.6) |
| Parity, n (%)* |  |  |
| Nulliparous | 720 (26.0) | 3413 (26.8) |
| Primiparous | 459 (16.6) | 2078 (16.3) |
| Multiparous | 1260 (45.4) | 5794 (45.5) |
| Unknown | 334 (12.0) | 1454 (11.4) |
| Age at first birth among parous women, n (%)* |  |  |
| <18 years old | 52 (3.0) | 273 (3.5) |
| 18-24 years old | 557 (32.4) | 2867 (36.4) |
| 25-29 years old | 637 (37.1) | 2766 (35.1) |
| 30-34 years old | 364 (21.2) | 1464 (18.6) |
| 35-39 years old | 93 (5.4) | 424 (5.4) |
| 40-44 years old | 12 (0.7) | 68 (0.9) |
| ≥45 years old | 1 (0.1) | 3 (0.0) |
| Unknown | 3 (0.2) | 7 (0.1) |
| Total length of breastfeeding among parous women, n (%)* |  |  |
| <12 months | 743 (43.2) | 3774 (47.9) |
| ≥12 months | 903 (52.5) | 3780 (48.0) |
| Unknown | 73 (4.2) | 318 (4.0) |
| Modifiable Factors |  |  |
| Body Mass Index, median [IQR]* | 24.7 [22.0, 29.0] | 25.0 [22.1, 29.1] |
| Oral contraceptive use, n (%) |  |  |
| Ever | 2469 (89.0) | 10927 (85.8) |
| Never | 304 (11.0) | 1803 (14.2) |
| Unknown | 0 (0.0) | 9 (0.1) |
| Tobacco use, n (%) |  |  |
| Ever | 1166 (42.0) | 5119 (40.2) |
| Never | 1600 (57.7) | 7591 (59.6) |
| Unknown | 7 (0.3) | 29 (0.2) |

\*These variables were collected at time of answering *My Health Overview* or *My Personal and Family Health History* surveys and may not accurately reflect the status at time of breast cancer diagnosis. Percentages may not sum to 100 due to rounding. Interquartile range, IQR.

Among those diagnosed with breast cancer, the most common age category at diagnosis was between 50 and 59 (Table 2). The majority of the breast cancers were of stages I and II (85.5%); 68.8% were ER positive and 43.7% were progesterone receptor positive (Table 2).

**Table 2.** Breast cancer characteristics of study participants who had infectious mononucleosis (IM) between ages 10 and 22 and those who never developed IM.

|  | Had IM | Never developed IM |
| --- | --- | --- |
|  | 2773 | 12739 |
| Breast cancer, n (%) |  |  |
| Yes | 311 (11.2) | 1686 (13.2) |
| No | 2462 (88.8) | 11053 (86.8) |
| Among breast cancer cases |  |  |
| Age at diagnosis, n (%) |  |  |
| <40 years old | 52 (16.7) | 248 (14.7) |
| 40-49 years old | 95 (30.5) | 546 (32.4) |
| 50-59 years old | 111 (35.7) | 590 (35.0) |
| 60-69 years old | 47 (15.1) | 266 (15.8) |
| ≥70 years old | 6 (1.9) | 36 (2.1) |
| Stage, n (%) |  |  |
| Stage I | 130 (41.8) | 743 (44.1) |
| Stage II | 131 (42.1) | 704 (41.8) |
| Stage III | 50 (16.1) | 239 (14.2) |
| Estrogen receptor status, n (%) |  |  |
| Positive | 215 (69.1) | 1159 (68.7) |
| Negative | 62 (19.9) | 312 (18.5) |
| Unknown | 34 (10.9) | 215 (12.8) |
| Progesterone receptor status, n (%) |  |  |
| Positive | 149 (47.9) | 723 (42.9) |
| Negative | 91 (29.3) | 523 (31.0) |
| Unknown | 71 (22.8) | 440 (26.1) |

Percentages may not sum to 100 due to rounding.

Women who had IM between the ages of 10 and 22 were less likely to develop breast cancer than those who did not develop IM after controlling for ethnicity, family history, age at menarche, oral contraceptive use, tobacco use, and birthplace (adjusted OR=0.82, 95% CI 0.72-0.93; Model 1). This association remained significant after additionally controlling for parity, age at first birth, BMI, and breast biopsy (adjusted OR=0.83, 95% CI 0.71-0.96; Model 2; Table 3A1). Our findings were unaltered when restricted to women born in the United States/Canada (Model 2: adjusted OR= 0.82, 95% CI 0.71-0.96; Table 3A2). When comparing women whose IM was diagnosed using a laboratory blood test versus those who did not develop IM, the findings were similar for Model 1 (adjusted OR=0.79, 95% CI 0.67-0.94) but was attenuated in Model 2 (adjusted OR= 0.88, 95% CI 0.72-1.07; Table 3A3).

**Table 3.** Association of infectious mononucleosis (IM) between ages 10 and 22 and breast cancer using logistic regression models to estimate odds ratios (ORs) and 95% confidence intervals (CIs).

|  |  |
| --- | --- |
| A1. IM and breast cancer |  |
| Cases (IM yes/no) / Controls (IM yes/no), n | (311/2462) / (1686/11053) |
| Crude | 0.83 (0.73-0.94) |
| Model 1 | 0.82 (0.72-0.93) |
| Model 2 | 0.83 (0.71-0.96) |
| A2. IM and breast cancer among women born in the United States/Canada |  |
| Cases (IM yes/no) / Controls (IM yes/no), n | (297/1595) / (2396/10550) |
| Crude | 0.82 (0.72-0.93) |
| Model 1 | 0.81 (0.71-0.92) |
| Model 2 | 0.82 (0.71-0.96) |
| A3. IM and breast cancer among women whose IM were diagnosed by a laboratory blood test |  |
| Cases (IM yes/no) / Controls (IM yes/no), n | (160/1686) / (1313/11053) |
| Crude | 0.80 (0.67-0.95) |
| Model 1 | 0.79 (0.67-0.94) |
| Model 2 | 0.88 (0.72-1.07) |
| B1. IM and breast cancer at <50 years old |  |
| Cases (IM yes/no) / Controls (IM yes/no), n | (147/794) / (2462/11053) |
| Crude | 0.83 (0.69-0.99) |
| Model 1 | 0.83 (0.69-0.99) |
| Model 2 | 0.82 (0.67-0.998) |
| B2. IM and breast cancer at >50 years old |  |
| Cases (IM yes/no) / Controls (IM yes/no), n | (164/892) / (2462/11053) |
| Crude | 0.83 (0.69-0.98) |
| Model 1 | 0.82 (0.69-0.97) |
| Model 2 | 0.83 (0.69-1.00) |
| C1. IM and estrogen receptor positive breast cancer |  |
| Cases (IM yes/no) / Controls (IM yes/no), n | (215/1159) / (2462/11053) |
| Crude | 0.83 (0.71-0.97) |
| Model 1 | 0.83 (0.71-0.97) |
| Model 2 | 0.84 (0.71-0.997) |
| C2. IM and estrogen receptor negative breast cancer |  |
| Cases (IM yes/no) / Controls (IM yes/no), n | (62/312) / (2462/11053) |
| Crude | 0.89 (0.67-1.17) |
| Model 1 | 0.87 (0.65-1.14) |
| Model 2 | 0.88 (0.65-1.16) |

Model 1 adjusted for ethnicity, family history, age at menarche, oral contraceptive use, tobacco use, and birthplace. Model 2 adjusted for the co-variables in Model 1 as well as parity, age at first birth, BMI, and breast biopsy.

The risk of developing invasive breast cancer by age of 50 was slightly lower in women who had IM versus those who did not develop IM (Model 2: adjusted OR=0.82, 95% CI 0.67-0.998; Table 3B1). Similar results were observed regarding the risk of developing invasive breast cancer after 50 years old between women who had IM versus those that did not develop IM (Model 2: adjusted OR=0.83, 95% CI 0.69-1.00; Table 3B2). When stratified by ER status, women who had IM were less likely to develop ER positive breast cancer compared to those who did not develop IM (Model 2: adjusted OR=0.84, 95% CI 0.71-0.997; Table 3C1). There was no association between IM and ER negative breast cancer (Model 2: adjusted OR=0.88, 95% CI 0.65-1.16; Table 3C2).

## Discussion

The link between EBV and breast cancer is controversial. The role of IM in the etiology of breast cancer remains unclear. Meta-analyses indicate that EBV is associated with increased breast cancer risk [32–34]. However, the studies included in the meta-analyses were not designed to elucidate whether EBV infection plays a role in the etiology of breast cancer. Population-based studies concluded null associations between IM and breast cancer [36–38]. We previously reported that *ex vivo* EBV infection of primary mammary epithelial cells, but not of established cancer cells, can lead to transcriptional re-programming and malignant transformation in organoid assays as well as to formation of breast cancers in a mouse model [20]. Therefore, we specifically designed this study using data from the HOW Study® to address whether the developing IM at the time of adolescent breast development when the population of mammary epithelial cells is expanding had an impact on later breast cancer development. In the HOW Study®, women who developed IM between the ages of 10 and 22 had a lower risk of developing breast cancer, particularly ER positive subtype, than those who did not develop IM. Our findings when taken together with previous population-based studies [36–38] suggest that an EBV vaccine is unlikely have an impact on reducing breast cancer risk.

There are several potential explanations for the discordance between pre-clinical studies [20,21] and the findings of this study: (1) The HOW Study® data did not allow us to address whether EBV infection at any age, including early childhood and asymptomatic exposures, modulates BC risk; (2) Most of the HOW Study® participants were white women. Thus, this study had insufficient power to address risk in women of African and South-East Asian descent, i.e. women from geographical areas where EBV-associated malignancies, triple-negative breast cancer, and early-onset breast cancer are more prevalent; (3) It is possible that EBV infection in humans does not result in effective infection of mammary epithelial cells even though EBV has been found to be excreted with breast milk [40]; and (4) It is also possible that EBV infected human mammary epithelial cells are effectively cleared by the immune system.

Previous population-based studies concluded a lack of association between IM and breast cancer [36–38]. The discordant findings between those studies and ours may be attributed to the difference in rate of IM cases. The demographics of the Nurses’ Health Studies is similar to the HOW Study®—college educated, white females born or residing in the United States and Canada—however the number of women diagnosed with IM between ages ≤15 to 24 (15.2%) in the Nurses’ Health Studies [37] was lower than our study (17.9%). Similarly, the rate of IM in Yasui *et al* was also lower than our study (31 had IM between ages 10 and 24 versus 972 never had IM; 3.2%) possibly due to their unique female population recruited in King County, Washington, United States. The IM rate among Swedish and Danish females in the study by Hjalgrim *et al* [38] was not reported. Nevertheless, our findings remained robust when restricted to laboratory-confirmed IM diagnosis. Age at breast cancer (<50 or ≥50 years old) also did not alter the results.

The strengths of our study include investigating the association of IM and breast cancer in a newly established, large research population. The HOW Study® collected exposure information about IM (age and method of diagnosis) and breast cancer risk factors. Limitations of our study include self-reported IM and self-reported method of IM diagnosis. We were unable to control for menopausal status, BMI, and alcohol intake at time of breast cancer diagnosis because those exposures were not specifically asked in the surveys. We made an effort to control for BMI at time of survey in our multivariable Model 2. The high degree of similarity between Models 1 and 2 presented in Table 3 confirms there is minimal confounding for the association between IM and breast cancer [37].

## Conclusion

Women in the HOW Study® who had IM between the ages of 10 and 22 were less likely to develop breast cancer, particularly of the ER positive subtype, compared to women who never developed IM. Our data do not support the development of an EBV vaccine for breast cancer prevention in the United States or Canada.

## Data Availability

The Health of Women (HOW) Study data that support the findings of this study are available from Dr. Susan Love Research Foundation but restrictions apply to the availability of these data, which were used under license for the current study, and so are not publicly available. Data access and policy details can be accessed at https://loveresearcharmy.org/researchers/faqs.

## Declarations

### Ethics approval and consent to participate

The Health of Women (HOW) Study® protocol was approved by the Western Institutional Review Board. Participants provided written consent.

### Consent for publication

Not applicable.

### Availability of data and materials

The Health of Women (HOW) Study® data that support the findings of this study are available from Dr. Susan Love Research Foundation but restrictions apply to the availability of these data, which were used under license for the current study, and so are not publicly available. Data access and policy details can be accessed at https://loveresearcharmy.org/researchers/faqs.

### Competing interests

All authors have no conflict of interest to disclose.

### Funding

GMW is supported by the AVON Foundation, the Breast Cancer Research Foundation, and the Ludwig Center at Harvard Medical School.

### Authors’ contributions

Yujing J Heng: Formal analysis, writing - review and editing; Susan Love: Resources, writing – review and editing; Jessica Clague DeHart: Data curation, writing – review and editing; Joyce D. Fingeroth: Conceptualization, writing – review and editing; Gerburg M. Wulf: Conceptualization, writing – review and editing

## Acknowledgements

We thank the staff and participants of the HOW Study ® and the Dr. Susan Love Research Foundation for providing this dataset. We thank Leah Eshraghi and Katherine M. Peterson for reading this manuscript. The authors assume full responsibility for analyses and interpretation of these data.

## Notes

### Competing Interest Statement

The authors have declared no competing interest.

### Author Declarations

The Health of Women (HOW) Study protocol was approved by the Western Institutional Review Board. Participants provided written consent.

